# Resurgence of Large-Scale Influenza A (H1N1) Outbreaks: Modeling the Interplay of Transmission, Loss of Immunity, and Vaccination

**DOI:** 10.1101/2025.06.21.25329874

**Authors:** Chanaka Kottegoda, Claudia T. Codeço, Claudio J. Struchiner, Lucas M. Stolerman

**Affiliations:** Department of Mathematics and Physics, Marshall University, Huntington, WV, USA; Programa de Computação Científica, Fundação Oswaldo Cruz, Rio de Janeiro, Brazil; School of Applied Mathematics (EMAp FGV), Brazil; Department of Mathematics, Oklahoma State University, Stillwater, OK, USA

**Keywords:** A(H1N1) influenza, Resurgent outbreaks, Mechanistic modeling

## Abstract

Influenza is a highly transmissible respiratory virus that can cause devastating pandemics. The last influenza pandemic, caused by a strain of A(H1N1) virus, resulted in millions of cases and hundreds of thousands of deaths worldwide. After the initial outbreaks, many regions experienced subsequent waves driven by environmental conditions, waning immunity, and other factors. More dramatically, several locations saw large resurgent outbreaks, yet the mechanisms behind these severe resurgences remain poorly understood. Here we investigate the dynamics of these large-scale H1N1 outbreaks using a mechanistic model with time-varying rates of infection, loss of immunity, and vaccination. In particular, we explore two mathematical functions to model dynamic loss of immunity, capturing how viral evolution may reduce the duration of immune protection. Numerical simulations reveal regimes that influence resurgence timing, and the model reproduces key features of H1N1 flu case data. We also fit the model to data from nine locations, demonstrating its ability to capture resurgence patterns and track associated changes in infection and immunity-loss rates. Our findings highlight the complex interplay among environmental drivers, population immunity, and viral mutation, contributing to a better understanding of large-scale H1N1 dynamics in the post-pandemic era.

## Introduction

A new strain of influenza A(H1N1) virus emerged in the spring of 2009 in central Mexico and rapidly spread worldwide [1]. The Centers for Disease Control and Prevention (CDC) reported the first US cases in April 2009, and the World Health Organization (WHO) declared a pandemic on June 11, 2009 [2]. From April 2009 to April 2010, the CDC reported 60.8 million cases, 274,304 hospitalizations, and 12,469 deaths in the United States alone [3]. Globally, many countries – including Brazil, China, Australia, and the United Kingdom – experienced significant surges between 2009 and 2010 [4, 5, 6, 7, 8]. The 2009 H1N1 virus has continued to circulate, causing subsequent outbreaks [9, 10].

Large H1N1 outbreaks have occurred worldwide in the post-pandemic era, often reaching incidence levels comparable to those of the initial 2009 pandemic. For instance, an estimated 30 million influenza illnesses, 13 million medical visits, 347,000 hospitalizations, and 38,000 deaths occurred in the United States during the 2013–2014 influenza season, when 2009 H1N1 viruses were the predominant circulating strains, primarily affecting young and middle-aged adults [11, 12, 13]. In 2016, Brazil experienced a severe outbreak with more than a thousand confirmed deaths [14]. Iran similarly faced a substantial resurgence during the 2015–2016 season, with elevated case counts and mortality [15, 16]. Similar spikes were reported in Turkey (2016), South Africa (2011), Croatia (2011), among other regions [17, 18, 19].

The mechanisms underlying sustained H1N1 transmission in the post-pandemic period are understood to an extent. During the 2009 outbreaks, susceptible populations were not fully depleted, allowing for subsequent waves of infection. As the number of susceptible individuals declined below critical thresholds, outbreaks became less frequent, resulting in “skip-and-resurgence” patterns [20]. Complex oscillations in influenza epidemics are thought to arise from a combination of epidemiological, immunological (humoral), and seasonal drivers, many of which have been successfully incorporated into mathematical models. Immunity acquired through natural infection wanes over time, while cellular immunity might last longer and play a role in the final dynamics [21]. Seasonal variations in transmissibility drive periodic increases in transmission [22, 23, 24, 25]. Poor vaccination coverage, suboptimal vaccine efficacy, and demographic changes further contribute to sustaining outbreaks [26, 27, 28]. Antigenic changes shape outbreak dynamics by rendering previously immune individuals susceptible to emerging strains [29, 30, 31].

The coupled roles of immunity, infection, and vaccination have been examined across a range of modeling frameworks. Recent work has introduced immuno-epidemiological models that explicitly incorporate immunity and infection ages to characterize epidemic dynamics [32, 33]. Mechanistic approaches have further investigated how waning immunity can shape viral evolution, demonstrating that immune-escape variants may arise dynamically through feedbacks between population-level immunity and viral adaptation [34, 35]. Other studies have shown that distinct epidemiological patterns can emerge from differences in the duration of vaccine-acquired versus infection-acquired immunity [36], and that models with multiple partial-immunity classes are capable of generating sustained epidemic waves [37]. Extensions incorporating graded susceptibility have yielded explicit conditions on vaccine supply required to maintain disease-free states [38], while frameworks allowing immunity boosting upon re-exposure highlight the critical role of immunity duration in shaping transmission dynamics [39]. In the specific context of post-pandemic influenza, a comprehensive global analysis by He et al. identified synchronization patterns and hypothesized that these arise from the combined effects of seasonal forcing, strain-specific immunity loss, and vaccination coverage [10]. Despite these advances, to our knowledge, no mechanistic model has yet been applied to specifically investigate the large resurgent H1N1 epidemics observed across multiple locations in the post-pandemic era.

Here, we address this gap by asking: how do transmission, loss of immunity, and vaccination shape the timing and magnitude of large resurgent H1N1 outbreaks? To investigate this question, we developed a mathematical model that incorporates time-varying rates for these three factors. In particular, we explore two functions to model dynamic immunity loss, capturing how viral evolution can shorten the duration of immunity in recovered individuals. Through numerical simulations, we identify parameter regimes that influence resurgence timing and show that our model reproduces key patterns observed in H1N1 time series data. By fitting the model to data from nine locations, we demonstrate that it captures the timing and key features of large resurgent outbreaks, reveal that transmission dynamics extend beyond simple seasonal forcing, and uncover location-specific patterns in immunity loss that suggest distinct antigenic drift trajectories across regions.

## Materials and Methods

### Description of epidemiological and demographic datasets

We identified locations that experienced large resurgent H1N1 outbreaks in the post-pandemic period by examining their weekly reported H1N1 case counts. Defining what constitutes a large resurgence is challenging and can be approached in multiple ways. Here, a large resurgent H1N1 outbreak is defined as one whose epidemic peak (maximum number of reported cases) exceeds 50% of the major peak observed during the 2009 pandemic. Major outbreaks in 2010 were not included and classified as late-pandemic activity rather than resurgences. Fig. 1 A) displays the nine selected locations/countries: São Paulo State in Brazil, Turkey, Iran, New Zealand, the U.S. Department of Health and Human Services (HHS) Regions 2 and 6, Colombia, South Africa, and Croatia. The HHS Region 2 includes the states of New Jersey and New York, as well as Puerto Rico and the U.S. Virgin Islands. The HHS Region 6 comprises the states of Arkansas, Louisiana, New Mexico, Oklahoma, and Texas.

**Figure 1:**
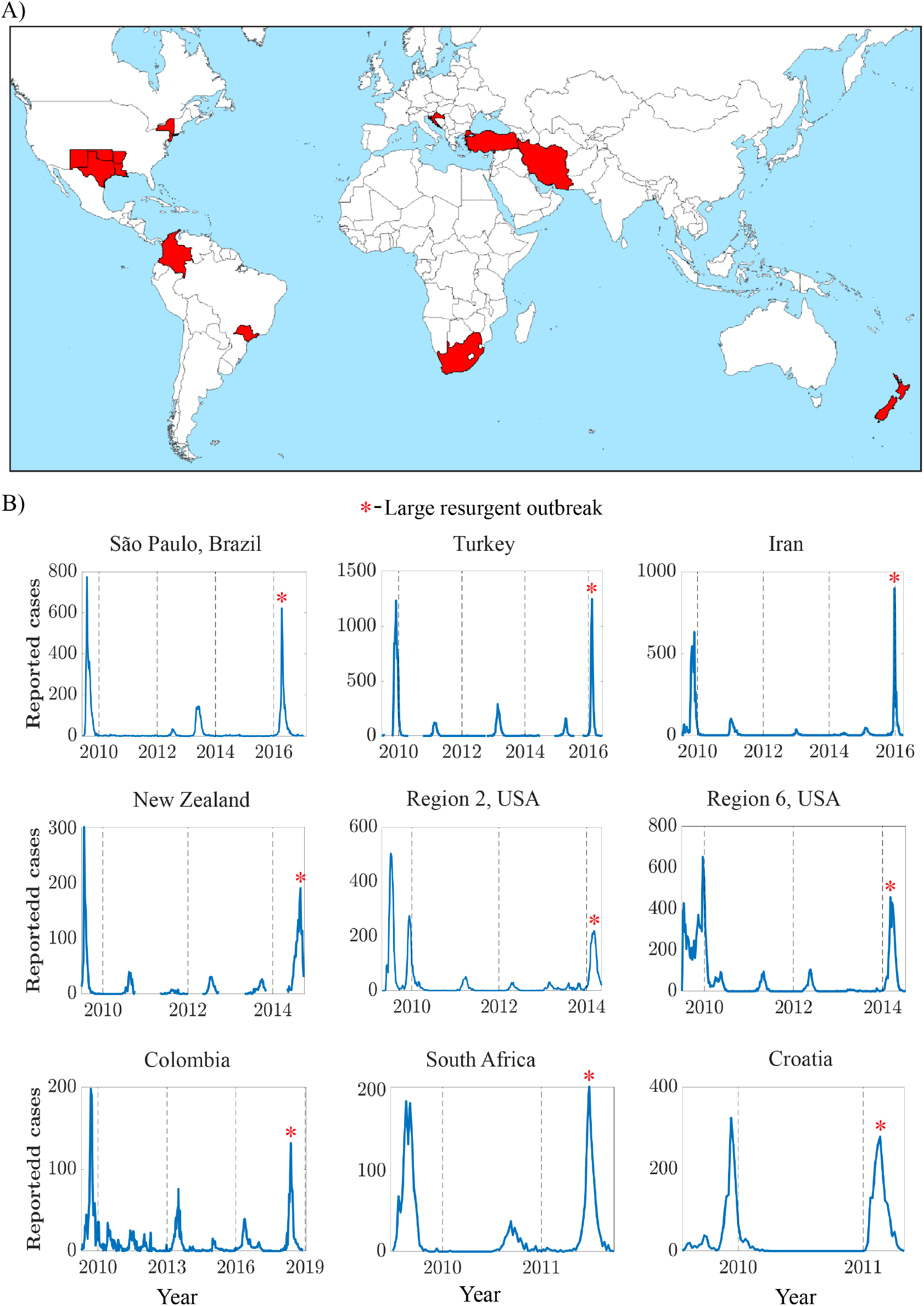
Weekly reported A(H1N1) influenza cases in selected regions. (A) Geographical locations of nine selected regions from eight countries that experienced a large resurgent H1N1 outbreak after the 2009 pandemic. (B) Weekly reported H1N1 cases in the selected regions, starting from the 2009 pandemic until the year of the large resurgent outbreak (marked by a red star). The visible gaps between the blue curves in New Zealand and Turkey indicate missing data.

Fig. 1 B) displays the time series of weekly reported H1N1 cases between 2009 and the first large resurgent outbreaks for each location. Some epidemiological datasets exhibited gaps in the data. In particular, New Zealand had 103 missing data points out of 273 weeks, Iran had eight missing data points out of 351 weeks, and Turkey had 69 missing data points out of 364 weeks. In this work, we did not perform data imputation or estimation for the missing values; only the available data were used in the model-fitting analysis. In Fig. 1 B), the large resurgent outbreaks are marked with a red star for each location. Epidemiological data from HHS Regions 2 and 6 were obtained from the Fluview interactive system [40]. H1N1 case counts for São Paulo were collected from the Brazilian Information System for Influenza Epidemiological Surveillance (SIVEP). The rest of the epidemiological data for other locations were obtained from FluNet [41], the World Health Organization’s (WHO) influenza surveillance system, which provides real-time, publicly accessible epidemiological data. For each location, we also collected population data from 2009 to the year of the large resurgence outbreak, sourced from Worldbank.org [42]. For datasets and further details, we refer the reader to the supplementary material.

### Mathematical Model

We developed an SIRS model with vaccination (SVIRS) to investigate the dynamics of large resurgent H1N1 outbreaks, as illustrated in Fig. 2 A) i). To maintain mathematical simplicity, the model omits age-structured dynamics, heterogeneous contact patterns, and exposure periods. Instead, we focus on the interplay of time-varying rates of infection, loss of immunity, and vaccination. The governing equations are given by the following system:

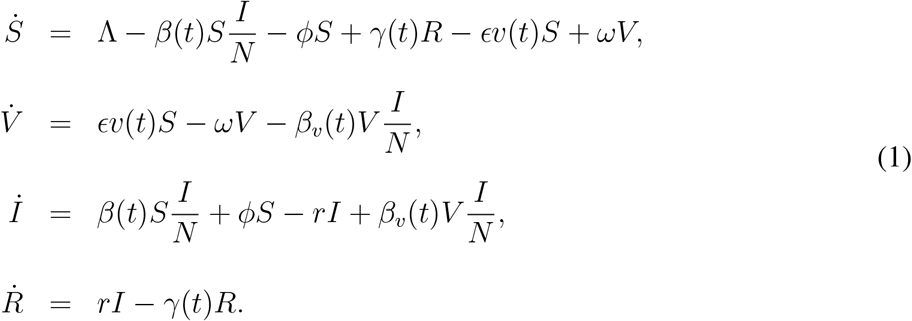

**Figure 2:**
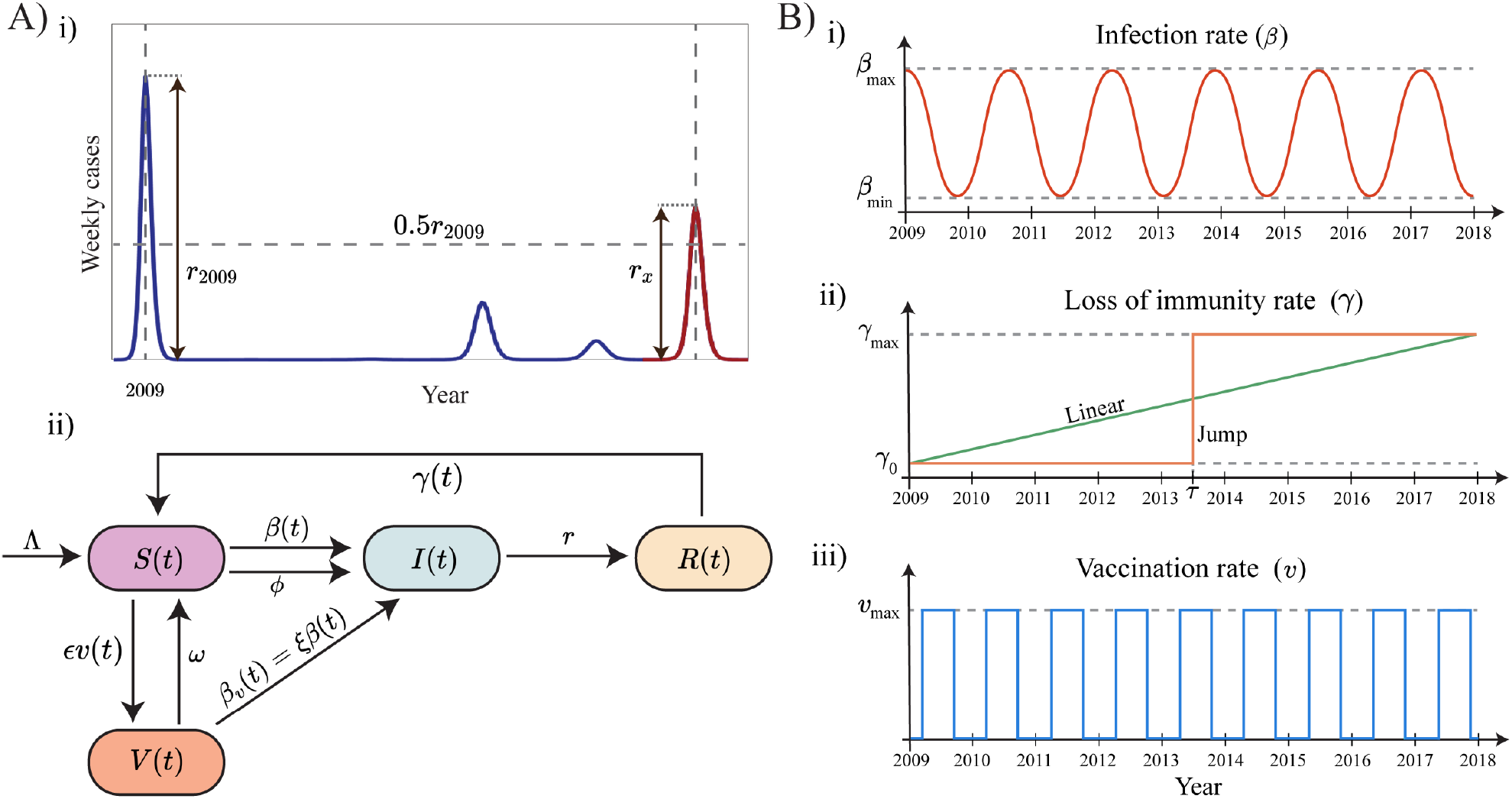
Large resurgent H1N1 outbreaks and our proposed SVIRS mathematical model. A) i) The resurgence of large H1N1 influenza outbreak in year *x* is defined by the condition *r*_*x*_ *>* 0.5*r*_2009_. Here *r*_2009_ represents the epidemic peak during the 2009 H1N1 pandemic, while *r*_*x*_ is the same quantity in year *x*. A) ii) A compartmental diagram of our SVIRS model. A population is divided into susceptible (S), vaccinated (V), infectious (I), and recovered individuals (R). The rates of Infection, loss of immunity, and vaccination are assumed to change over time. B) i) A sine function was used to approximate the periodic infection rate *β*(*t*), with minimum and maximum values *β*_min_ and *β*_max_. B) ii). Linear and Jump functions represent different effects of antigenic drift on the loss-of-immunity rate *γ*(*t*) that increases from *γ*_0_ to *γ*_max_. B) iii). A rectangular pulse function represented the vaccination rate, which increases to *v*_max_ during vaccination periods.

Here, *S, V, I*, and *R* represent the susceptible, vaccinated, infectious, and recovered individuals, respectively. Overdots denote derivatives with respect to time. The total population is given by *N* = *S* + *V* + *I* + *R*. The time-varying rates are assumed to reflect dynamic changes driven by complex biological factors, which are discussed in detail in the next subsection. Here, *β*(*t*) denotes the infection rate at which susceptible individuals become infectious. The parameter *γ*(*t*) represents the loss-of-immunity rate, and *v*(*t*) the vaccination rate. Additionally, *β*_*v*_(*t*) = *ξβ*(*t*) denotes the infection rate at which vaccinated individuals become infectious. The factor *ξ* between 0 and 1 represents the vaccine efficacy, where *ξ* = 0 corresponds to a vaccine that confers perfect protection. The recruitment rate Λ is constant and thus *N* (*t*) satisfies the differential equation 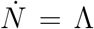, leading to *N* (*t*) = Λ*t* + *N*_0_, where *N*_0_ denotes the total initial population. This linear trend provides a suitable approximation for the complex demographic evolution observed in the selected locations during the initial years following the 2009 H1N1 pandemic (see supplementary material). Natural recovery rates are represented by the parameter *r*, while *ω* describes the rate at which vaccine-induced immunity wanes. Finally, *ϵ* and *ϕ* represent the vaccine effectiveness and a small importation rate, respectively. We list all fixed (time-independent) parameters in Table 1 (see supplementary material for details).

**Table 1:** Time-independent model parameters: description and values.

| Parameter | Description | Value |
| --- | --- | --- |
| $\Lambda$ | Recruitment rate into the susceptible class | Location-specific; estimated from demographic data (see supplementary material) |
| $r$ | Natural recovery rate | 1.17 week<sup>-1</sup> [43] |
| $\phi$ | Importation rate | 10<sup>-6</sup> week<sup>-1</sup> (one imported case per million susceptibles per week) |
| $\epsilon$ | Vaccine effectiveness | 0.7 [44] |
| $\omega$ | Waning rate of vaccine-induced immunity | 0.019 week<sup>-1</sup> [45] |

#### Model assumptions for time-varying rates

##### Periodic infection rates

Influenza transmission depends on multiple environmental and social factors with seasonal components, including humidity [46, 47, 48], human mobility [49, 50, 51], and school terms [52, 53]. We adopt a time-varying periodic infection rate, *β*(*t*), given by the following expression:

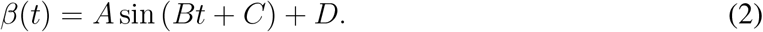

The parameters *B* and *C* correspond to the angular frequency and phase shift of the sine function, while *A* and *D* represent its amplitude and vertical shift. To parameterize *β*(*t*) in terms of its maximum and minimum values *β*_max_ and *β*_min_, we can simply write 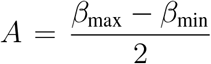 and 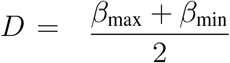. Fig. 2 B) i) illustrates an example of a periodic *β*(*t*), representing periodic changes in the infection rate.

##### Loss-of-immunity rates

The influenza virus evolves through two main processes: antigenic drift and antigenic shift [54]. While antigenic shift can lead to the abrupt emergence of novel subtypes – such as the 2009 H1N1 pandemic strain – our modeling approach focuses on antigenic drift, the gradual accumulation of mutations that help the virus evade host immune responses [55, 56, 57].

We represent drift using two formulations: a linear model, which assumes continuous immune escape over time [58], and a jump model, which describes a punctuated antigenic change [59]. This latter mechanism is consistent with evidence that a single mutation at a key site, such as on the hemagglutinin (HA) protein, can render the virus antigenically distinct [60]. Mathematically, we consider the following functions:

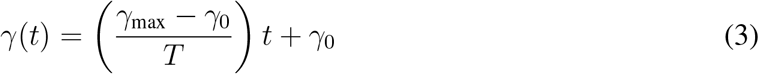

and

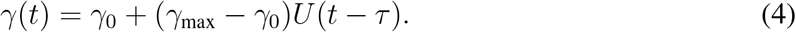

For the linear model (Eq. (3)), *γ*_0_ and *γ*_max_ denote the baseline and maximum rates of loss of immunity, respectively, with *T* the interval over which *γ* rises from *γ*_0_ to *γ*_max_. We also set *T* in weeks based on the time between the initial outbreak and the year of the large resurgence in each location. Throughout this paper, we fix *γ*_0_ = 3.2 *×* 10^−3^ week^−1^ (see supplementary material for details). In the jump model (Eq. (4)), *γ*_max_ again defines the post-jump maximum loss rate, and *τ* specifies the time of the discrete increase. The term *U* (*t* − *τ* ) represents a step function assuming the values 0 if *t < τ* and 1 if *t* ≥ *τ* . In what follows, we refer to the *linear immunity decay model* whenever the SVIRS model (Eqs. (1)) is equipped with the linear function given by Eq. (3). Similarly, the *jump immunity decay model* refers to the SVIRS model with *γ*(*t*) given by Eq. (4).

##### Vaccination rates

The frequent antigenic changes prompt re-evaluations of influenza vaccines, which are typically administered yearly before the flu season. For example, in Brazil, vaccines are offered to high-risk groups through annual campaigns between April and May, a period that generally precedes the typical flu seasons [61]. We adopted a vaccination rate *ϵv*(*t*) where *v*(*t*) is a rectangular pulse that achieves the maximum level *v*_max_ during the vaccination period for a given location (see Table S2 in the supplementary material). Fig. 2 B) iii) illustrates the format of *v*(*t*).

## Numerical Simulations

We performed numerical simulations to explore parametric regimes that influence the timing of large resurgent outbreaks and to evaluate our model’s ability to capture the overall characteristics of reported H1N1 case time series. Fig. 3 displays four sets of simulations conducted under different vaccine efficacy factors (*ξ*), considering the linear immunity decay model (Eq. (3)). Results for *ξ* = 0, 0.3, 0.6, and 0.9 are shown in panels A)–D). The colormaps depict the time interval (in years) between large outbreak peaks as a function of the minimum immunity period 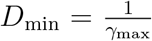 and the maximum infection rate *β*_max_. For all colormaps, the time interval between large outbreaks tends to increase as the immunity period lengthens. Lower vaccine efficacy (i.e., larger *ξ* values) resulted in shorter time intervals between large outbreaks. That can be observed as colormaps become darker as *ξ* increases. We also display sample model trajectories for selected points in the colormaps, which exhibit decreasing time intervals between large outbreaks as *ξ* increases. The leftmost time series of each colormap shows the modeled weekly H1N1 cases for low *β*_max_ and *D*_min_. The resulting time interval between the first and large resurgent outbreaks decreases from 4 years to 1 year, as *ξ* changes from 0 to 0.6. Within each colormap, longer immunity periods tend to delay the occurrence of large resurgent outbreaks. For instance, in Fig. 3 A), three sample trajectories are highlighted. At the lowest *D*_min_ (approximately 8 weeks [62]), a large resurgent outbreak occurs 4 years after the initial outbreak. In contrast, at the highest *D*_min_ value (approximately 30 weeks), no large resurgent outbreak is observed within ten years, as indicated by the dark blue region marked (NR).

**Figure 3:**
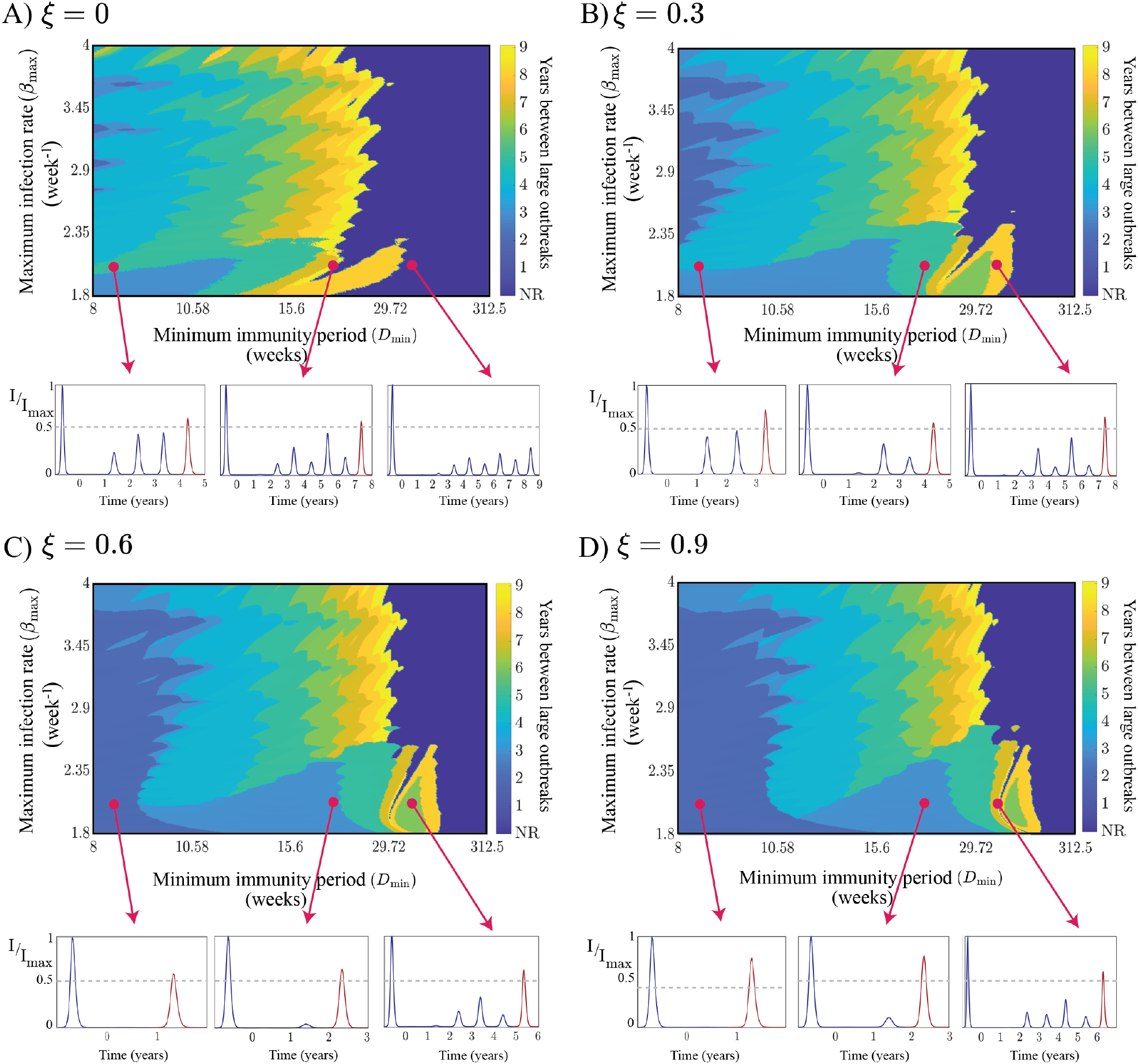
Years between large outbreak peaks along with sample trajectories (linear immunity decay model). Panels (A)–(D) show colormaps of the time interval (in years) between large outbreak peaks for increasing values of *ξ* (0, 0.3, 0.6, 0.9), corresponding to decreasing vaccine efficacy. Dark blue regions labeled “NR” indicate no large outbreaks within the past ten years. Beneath each colormap are sample trajectories of the normalized infectious population. Gray dashed lines indicate the threshold used to define large resurgent outbreaks (red colored). Parameter values were taken from Table 1. Initial population *N*_2009_ = 42, 075, 716 and recruitment rate Λ = 137.9351 week^−1^ were taken from São Paulo, Brazil. Other fixed parameters include *β*_min_ = 1 week^−1^, *v*_max_ = 0.02 week^−1^, *γ*_0_ = 3.2 *×* 10^−3^ week^−1^, and *T* = 520 weeks (10 years). For the periodic infection rate, we set *B* = −0.1202 week^−1^ and *C* = −10.1360 to produce annual cycles.

Fig. 4 shows numerical simulations of the jump *γ* model (Eq. (4)). We set *τ* = 130 weeks (2.5 years), representing antigenic changes occurring 130 weeks after the initial outbreak, and *v*_max_ = 0.01 week^−1^ while all other parameters remain the same as in Fig. 3. The predominance of light blue colormap regions across panels A–D indicates immediate large resurgent outbreaks following the jump, while yellow regions highlight outbreaks delayed by several years. Sample trajectories below each colormap illustrate these varied dynamics; Notably, the middle time series in panels C and D exhibit resurgent outbreaks that slightly exceed the threshold (gray dashed line) approximately seven years after the initial wave.

**Figure 4:**
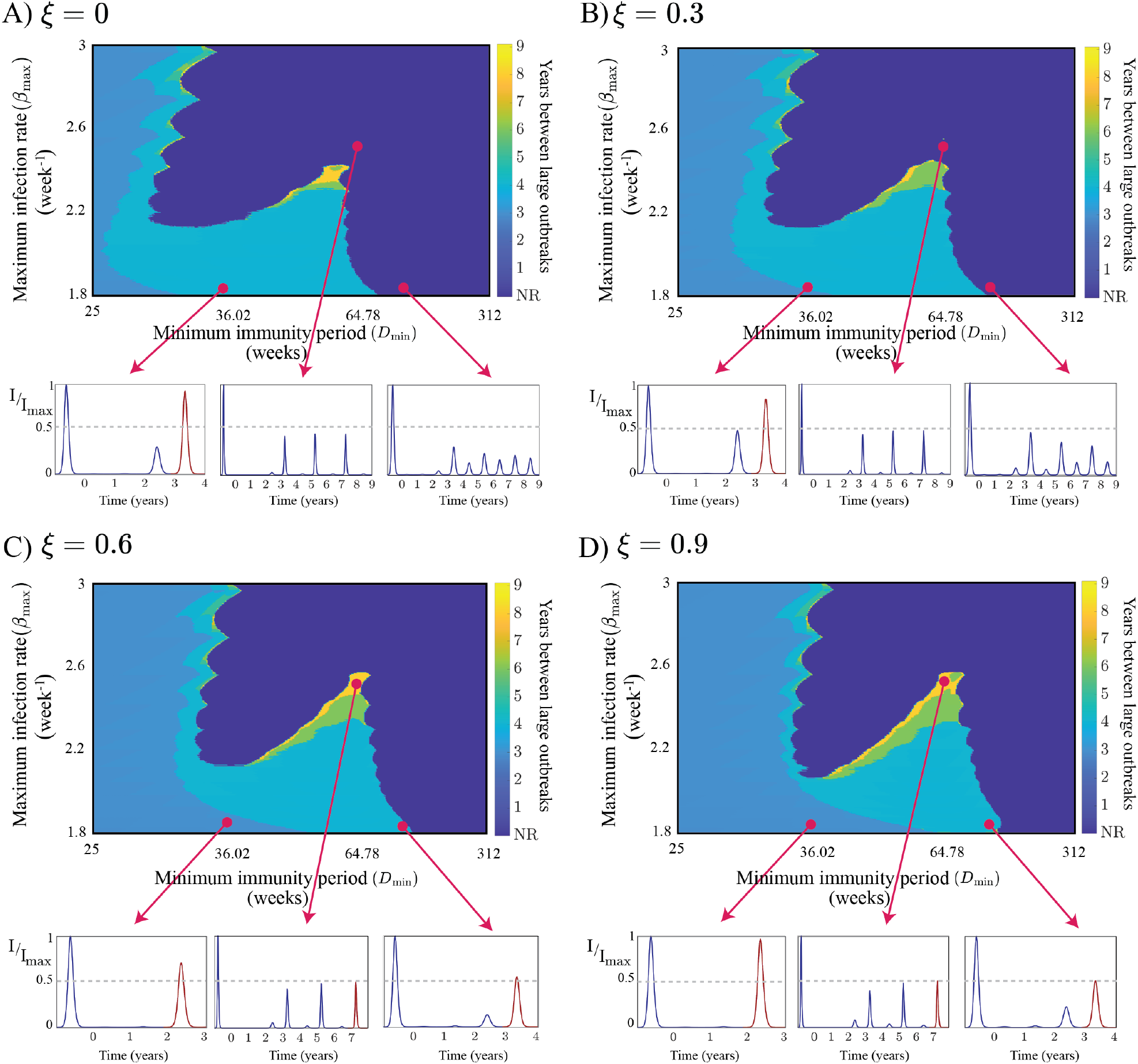
Years between large outbreak peaks along with sample trajectories (jump immunity decay model). Panels (A) - (D) exhibit four colormaps for increasing values of *ξ*, corresponding to decreasing vaccine efficacy levels. We considered four different *ξ* values (*ξ* = 0, 0.3, 0.6, and 0.9). Here, *ξ* = 0 represents a vaccine that confers perfect protection (panel A). Dark blue regions indicate no large resurgent outbreaks within ten years (labeled “NR”). Here, we set *τ* = 2.5 years and *v*_max_ = 0.01 week^−1^ while all other parameter values remain the same as in Fig. 3.

## Fitting the model to H1N1 data

We evaluated our model’s ability to fit H1N1 case data across the nine locations shown in Fig. 1 – São Paulo State in Brazil, Turkey, Iran, New Zealand, HHS Regions 2 and 6 in the US, Colombia, South Africa, and Croatia. These selected locations experienced a large resurgence of H1N1 outbreaks within a decade of the 2009 pandemic, a timeframe that captures the combined effects of waning immunity, cycles of higher transmission, and continued vaccination campaigns. For each location, we performed a least-squares optimization, fitting the simulated reported cases to the available case data.

### Optimization Procedure for Model Fitting

For the linear immunity decay model (Eqs. (1) and (3)), we estimated eight unknown parameters for each location. For simulations involving the jump *γ* model (Eq. (4)), we estimated a total of nine parameters. See Table 2 for the list of the estimated parameters for each model. Other parameters were fixed at the values listed in Table 1, along with a fixed baseline loss-of-immunity rate *γ*_0_ = 3.2 *×* 10^−3^ week^−1^ (see supplementary material). We also fixed *β*_min_ = 1 week^−1^ so that during off-peak seasons we have *β*(*t*) *< r*, ensuring natural epidemic decline in troughs, while allowing *β*_max_ to drive seasonal surges.

**Table 2:** List of the estimated parameters for each model. Parameter estimation involved eight unknowns per location for the linear immunity decay model (Eqs. (1) and (3)) and nine for the jump immunity decay model (Eqs. (1) and (4)).

| Parameter | Description | Immunity Decay Model |
| --- | --- | --- |
| $\beta_{\max}$ | Maximum infection rate | Linear and Jump |
| $\gamma_{\max}$ | Maximum rates of loss of immunity | Linear and Jump |
| $v_{\max}$ | Maximum vaccination rate | Linear and Jump |
| $\tau$ | Time where the jump occurs in the loss-of-immunity rate | Jump only |
| $B$ | Angular frequency of $\beta(t)$ function | Linear and Jump |
| $C$ | Phase shift of $\beta(t)$ function | Linear and Jump |
| $\xi$ | Vaccine efficacy | Linear and Jump |
| $I_0$ | Initial infectious population | Linear and Jump |
| $\rho$ | Reporting rate | Linear and Jump |

To obtain the best-fit curves, we performed grid searches over the model parameters within predefined lower and upper bounds. For the linear immunity decay model, four evenly spaced values were assigned to each of the eight parameters, yielding 4^8^ = 65,536 possible initial guesses. For each initial guess *θ*_*k*_, *k* = 1, 2, …, 4^8^, a least-squares optimal parameter value 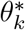 was obtained. Here, the objective function is the sum of squared residuals (SSR) given by 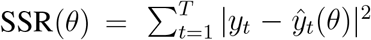 where *y*_*t*_ and *ŷ*_*t*_(*θ*) denote the observed data and model-predicted values, respectively, and *T* is the total number of weekly observations for the given location. For simplicity, we considered *ŷ*_*t*_(*θ*) = *ρI*_*t*_(*θ*) under the assumption that modeled reported cases are a fraction of prevalence. Among all 4^8^ resulting optimal parameters 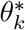, we chose *θ*_opt_ as the one yielding the minimum SSR across iterations, i.e., 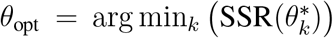. See Algorithm (1) in the supplementary material for further details. Optimization was performed using MATLAB’s `lsqcurvefit` function. The system of ODEs was numerically integrated in MATLAB using the `ode45` function, which employs a fourth-order Runge–Kutta method. A similar procedure was applied to the jump *γ* model, where three equally spaced initial guesses were used per parameter, resulting in 3^9^ (19, 683) simulations. As this was a sparser search compared to the 4^8^ initial guesses used for the linear model, we conducted an additional 10,000 simulations using randomly sampled starting points to enhance the search for the global minimum.

To find the Uncertainty bounds for the best-fit curve for each location, we used a parametric bootstrapping approach as described in [63]. First, we computed the cumulative incidence curve *F* (*t*) defined by 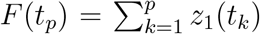 where *z*_1_(*t*_*k*_) represents the number of cases in week *t*_*k*_. For all of our selected regions, the empirical variance of weekly cases is greater than the mean, indicating overdispersion. Hence, we added a negative binomial distribution to the best-fit curve to incorporate noise in the data. Precisely, new simulated incidence data were generated as *y*_*t*_ ∼ NB(*r, p*) where parameters *r* and *p* were derived from the empirical mean (*µ*_*t*_) and variance 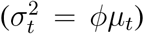 using 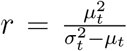 and 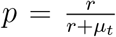. For each location, a total of 100 realizations were generated for the model’s best-fit using the negative binomial distribution. The model was subsequently refitted to every simulated dataset, with the best-fit set of parameters (*θ*_*opt*_) taken as the initial guess. For each of the 100 fits, we recorded the optimized parameter values and the associated residual sum of squares. From these replicates, 95% confidence intervals (CIs) for predicted incidence were computed as 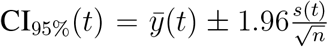 where 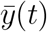 and *s*(*t*) denote the mean and standard deviation across all bootstrapped curves at time *t*.

### Best-fit curves and biological interpretation: linear immunity decay model

The best-fit curves for the linear *γ* model are presented in Fig. 5. Uncertainty bounds, shown in purple shades, were obtained as discussed above. Overall, the linear immunity decay model accurately captured the timing of most initial and large resurgent outbreaks. The closest fits occurred in South Africa, Croatia, and HHS Region 2 (USA). The model fits also showed reasonable agreement for São Paulo, New Zealand, and Iran, although sometimes over- or underestimating outbreak magnitudes. Remarkably, despite substantial gaps in the New Zealand data, the model recovered the large outbreaks and even inferred a surge around 2012 during the unreported interval. In contrast, mid-sized surges were less accurately captured in other locations. Model performance was slightly weaker in HHS Region 6 (USA), Turkey, and Colombia; notably, Colombia was the only location where the model failed to capture a clear resurgent wave, likely reflecting higher noise in the reported data. HHS Regions 2 and 6 in the United States experienced two pronounced H1N1 waves—in 2009 and again in 2010. Croatia followed a similar trajectory, with a modest preliminary wave occurring before its major 2009 outbreak. To account for these early peaks in 2009, we fitted the first outbreak to an *SIRS* model with constant rates of infection and loss of immunity. The resulting values of *S, I*, and *R*, together with *V* = 0, were then used as initial conditions for a second simulation using our SVIRS model, targeting the period between the 2009–2010 waves and the later large resurgence.

**Figure 5:**
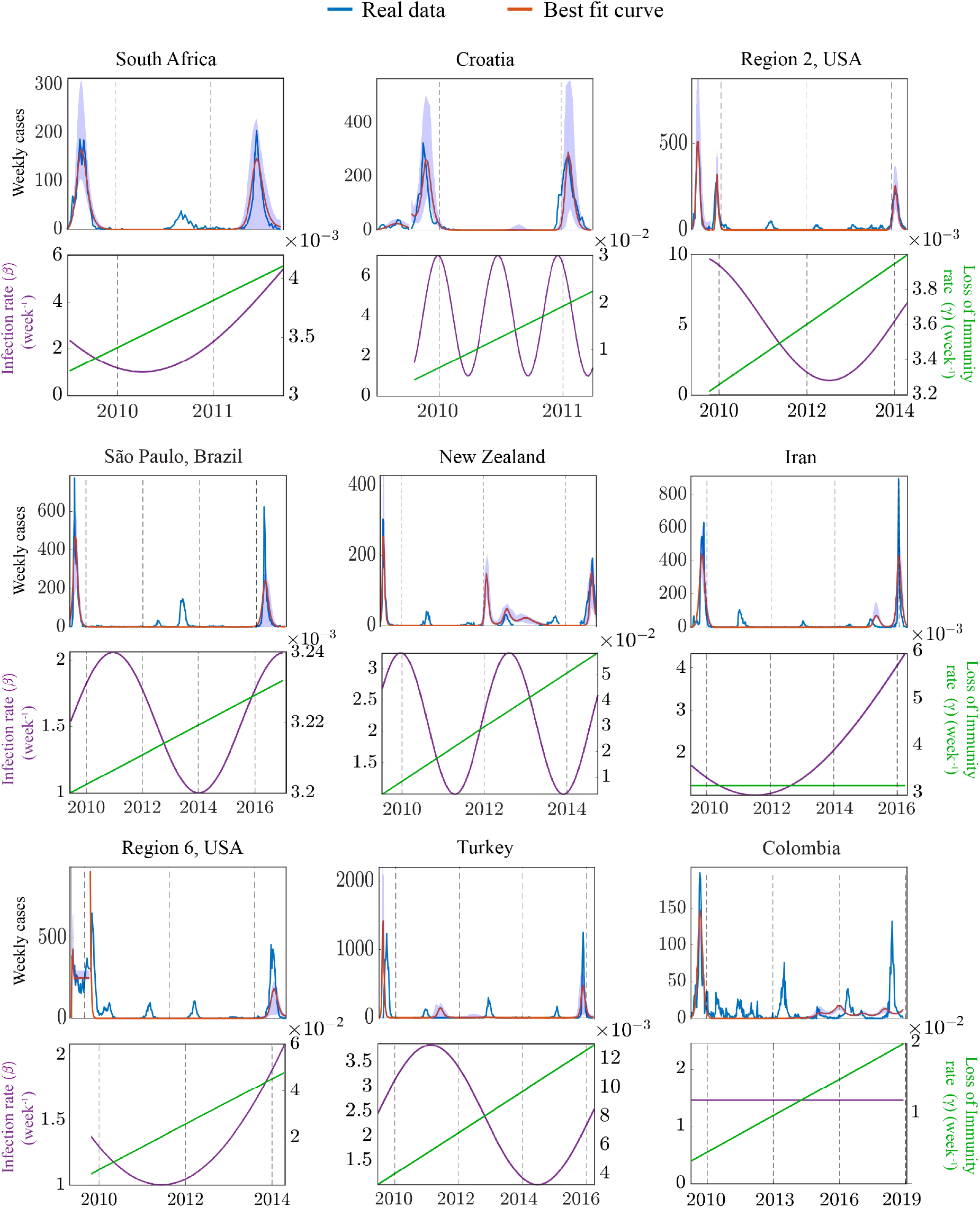
Best-fit curves and optimal time-varying rates (linear *γ* model). The top panels show observed (blue) and modeled (red) weekly H1N1 cases for nine locations, with shaded uncertainty bounds. Bottom panels display the fitted infection rate *β*(*t*) (purple) and linearly increasing immunity-loss rate *γ*(*t*) (green).

At the bottom of each best-fit panel in Fig. 5, the estimated rates of infection and loss of immunity illustrate the interplay between these two factors in shaping H1N1 dynamics up to the large resurgent outbreak. A flexible range of oscillations in the infection rate (dark blue) was observed, suggesting that complex transmission spikes arise from factors beyond seasonal influences. This implies that climate-driven oscillations alone may not fully explain the intricate temporal patterns of H1N1 outbreaks, particularly the resurgence events. For example, Croatia exhibited pronounced increases in infection rates during the winter flu season months (vertical dashed lines aligned with the beginning of each year), consistent with expected Northern Hemisphere seasonality. It also displayed a summer spike likely reflecting increased contact patterns due to tourism and summer activities. However, no outbreak was observed in the data nor in our best-fit solution, suggesting the susceptible pool was insufficient to sustain transmission despite elevated *β*(*t*). In contrast, South Africa did not show marked oscillations in *β*(*t*), but rather a gradual increase in transmission, possibly caused by the resumption of normal contact patterns following the pandemic year. An interesting decrease-followed-by-increase profile was found in HHS Regions 2 and 6 (USA), possibly reflecting a complex combination of environmental and behavioral factors: the initial decrease may correspond to residual post-pandemic behavioral caution, while the subsequent rise could indicate waning of both protective behaviors coupled with seasonal transmission drivers.

For locations where the model fits were less accurate, biological interpretations should be made with caution. Nonetheless, reasonable fits for São Paulo and New Zealand in the Southern Hemisphere exhibited oscillations that are unlikely to be explained by the same mechanisms driving Northern Hemisphere dynamics. In São Paulo, peaks of high transmissibility were estimated in 2011 and 2016, the latter coinciding with the resurgent outbreak. New Zealand exhibited a sharp increase in *β*(*t*) during 2012–2013, allowing the model to capture one of the minor waves observed in the data and, interestingly, projecting a larger wave in 2012– a period for which no surveillance data is available.

Loss-of-immunity rates *γ*(*t*) are plotted in green on a separate y-axis. In South Africa, *γ*(*t*) increased from the baseline value of 3.2 *×* 10^−3^ to approximately 4 *×* 10^−3^ week^−1^, corresponding to a decrease in the immunity period from 312 weeks (6 years) to 250 weeks (4.8 years)—a modest reduction consistent with gradual antigenic drift rather than a punctuated immune escape event. A similar pattern was observed for HHS Region 2. In contrast, a more aggressive decline in immune protection was estimated for Croatia, New Zealand, and HHS Region 6. For example, in Croatia, *γ*(*t*) reached levels of 2 *×* 10^−2^ week^−1^, indicating a decrease in the immunity period to approximately one year, possibly reflecting more rapid antigenic drift in the circulating strains in that region. Notably, Iran’s panel shows a virtually flat *γ*(*t*) – no appreciable increase in immunity loss –, yet the model still achieves a good fit. This suggests that our model can infer a possible absence of a significant antigenic drift over the considered period. All estimated parameters and confidence intervals can be found in the supplementary material (Tables S4 – S12).

### Best-fit curves and biological interpretation: jump immunity decay model

Fig. 6 presents a similar set of best-fit results for the jump immunity decay model. The model captured the overall timing and magnitude of large resurgent outbreaks in most locations. The bottom panels display the fitted rates of infection and loss of immunity. Interestingly, the jump immunity decay model enabled exploration of antigenic change timing. For instance, in South Africa, a jump was inferred between 2010 and 2011, reasonably close to the resurgent outbreak. In contrast, in São Paulo, the jump occurred closer to 2010, while the resurgent outbreak only took place in 2016. One possible interpretation is that antigenic changes preceding a large resurgent outbreak by several years may not immediately trigger resurgences if the susceptible pool has not yet been sufficiently rebuilt. However, the early jump timing may also reflect the model’s structural constraint of a single jump, effectively approximating a sustained elevated loss-of-immunity rate over the full period. In some locations, such as Croatia and HHS Region 2 (USA), the best-fit solutions exhibited jumps at the beginning of the time series, suggesting that a constant immunity decay model might better describe influenza dynamics in these regions. Biologically, this could represent scenarios where immunity loss remained stable over time due to potentially mild antigenic changes.

**Figure 6:**
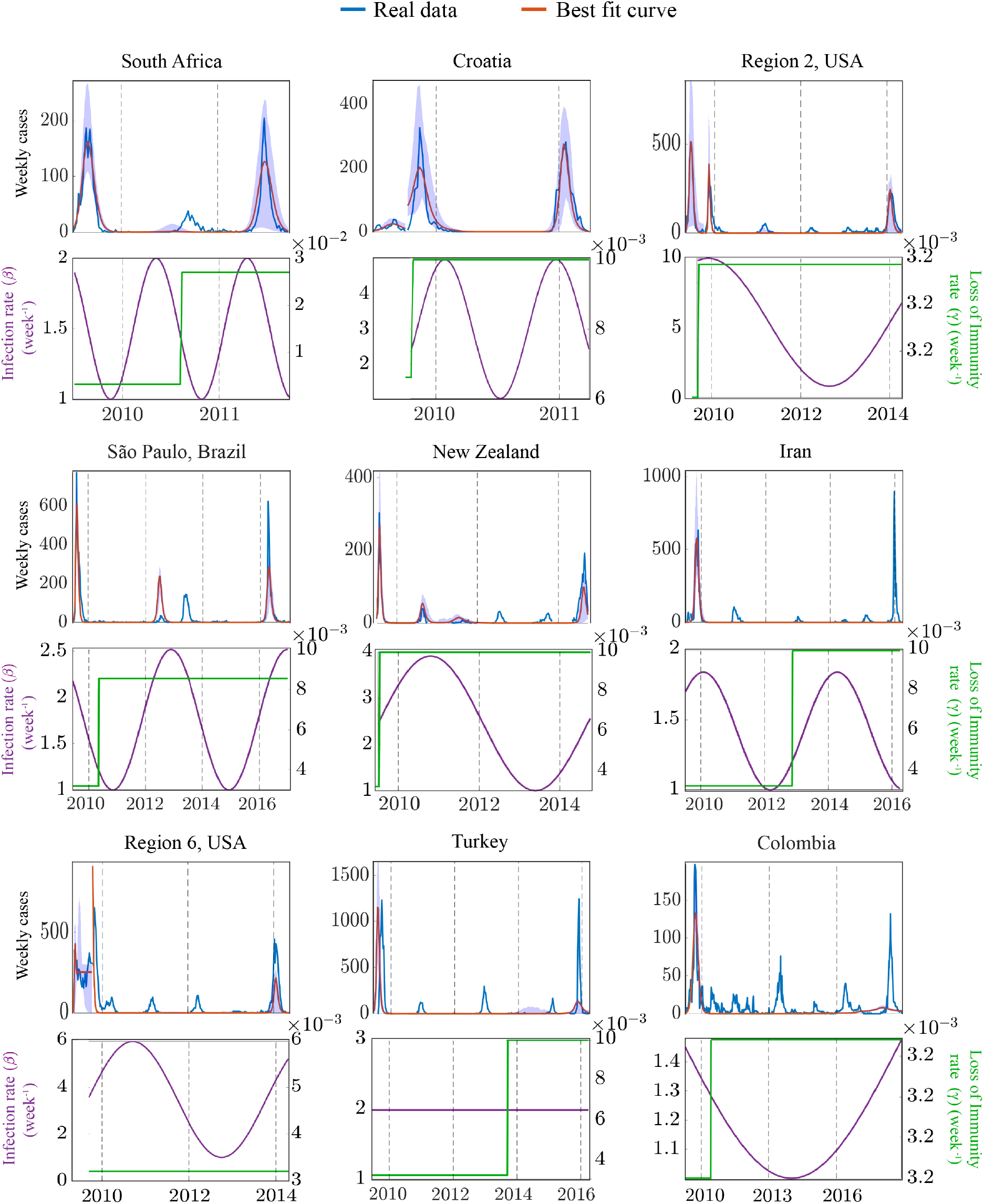
Best-fit curves and time-varying rates (jump immunity decay model). Top panels show observed (blue) and modeled (red) weekly H1N1 cases for nine locations, with shaded uncertainty bounds. Bottom panels display the fitted infection rate *β*(*t*) (purple) and jump immunity-loss rate *γ*(*t*) (green).

Oscillation periods of *β*(*t*) varied by location, as in the linear immunity decay model. Best-fit solutions with higher-frequency *β*(*t*) oscillations were observed in South Africa, Croatia, and São Paulo, while lower-frequency oscillations were found in HHS Regions 2 and 6 (USA). As in the linear immunity decay model, higher-frequency oscillations may reflect regions where transmission is modulated by multiple factors within a single year, such as climate variability and distinct waves of social activity, whereas lower-frequency oscillations suggest more stable transmission dynamics or a lack of pronounced seasonality. South Africa exhibited good fits under a broader range of oscillatory patterns and *γ* values than those obtained with the linear immunity decay model, suggesting that seasonal winter activity combined with punctuated antigenic drift may also explain the observed resurgent outbreak. Similarly, Croatia displayed pronounced winter peaks consistent with the linear model; however, the best-fit *β*(*t*) did not include the summer peak inferred from the linear model. For São Paulo, the best-fit solution showed an increase in *β*(*t*) between 2012 and 2014, capturing one of the smaller outbreaks—albeit with a slight overestimation of its magnitude—that preceded the major resurgence in 2016.

## Discussion

Large-scale resurgent H1N1 outbreaks have emerged worldwide in the post-pandemic era, often reaching incidence levels comparable to those observed during the 2009 pandemic. To investigate these resurgences, we developed a mechanistic SVIRS model with assumptions grounded in biological and demographic drivers. Infection rates were chosen to fluctuate periodically to reflect the impact of environmental and social factors on transmission. We modeled loss of immunity dynamically using linear and jump functions to represent different effects of antigenic drift. Vaccination rates were represented by a time-varying pulse to reflect targeted seasonal campaigns. We investigated how different parameter combinations influence outbreak dynamics and assessed model fits to historical H1N1 data across nine locations.

Using numerical simulations, we identified parameter regimes that influence the timing of large resurgent outbreaks. As shown in Fig. 3, longer immunity durations increased the interval between large peaks, while reduced vaccine efficacy shortened these intervals. We then fitted both linear and jump immunity decay models to historical H1N1 data from nine locations. Both models generally captured the timing of initial and resurgent outbreaks, though accuracy varied across locations. The closest fits occurred for South Africa, Croatia, and HHS Region 2 (USA), where the models effectively captured outbreak peaks. In contrast, fits for São Paulo, New Zealand, Iran, Turkey, and Colombia showed some discrepancies, particularly in capturing outbreak magnitudes or precisely aligning with observed peaks. The fitted infection-rate oscillations varied considerably, suggesting that additional drivers beyond seasonal forcing may contribute to resurgent outbreaks.

Two time-varying functions were proposed to model how waning immunity replenishes the susceptible pool. Previous work by Yaari et al. employed a more sophisticated version of the jump immunity decay model, incorporating multiple jumps corresponding to different influenza seasons to capture seasonal dynamics [64]. In contrast, our approach simplifies this idea by modeling immunity loss through a single jump or a linear increase. Biological evidence supports this simplification, as the H1N1pdm strain has remained antigenically stable in recent years [65]. To our knowledge, a linear function has not previously been used to model the effects of antigenic drift. It remains an open question whether using an evolution-informed approach—as in Du et al. [31]— would yield substantially different estimates for *γ*(*t*). In some scenarios, such as regions that experienced a single dominant antigenic change, we suspect that our simplified *γ*(*t*) functions may closely approximate more complex evolution-informed structures.

Our study has limitations. In some locations, the different model choices resulted in similar good fits. For example, for South Africa, the jump model yielded a substantially higher loss-of-immunity rate compared to the linear model, yet both achieved comparable fits to the data. Hence, the same outbreak dynamics could be explained either by gradual antigenic drift over time or by a punctuated antigenic change followed by rapid immune escape. Rather than selecting a single mechanism, we interpret these results as evidence that both scenarios remain plausible explanations for the observed resurgence, and distinguishing between them would require additional data such as viral sequence information or serological surveys. While our model captured key features of large resurgent outbreaks, the modest fit in some locations suggests that further refinement is needed to better capture epidemic dynamics. Specifically for locations with many missing data points, such as New Zealand, fitting our models to extended data using imputation techniques remains an interesting exercise for future studies. Our current framework does not account for multi-strain dynamics, which are known to influence influenza patterns and may contribute to discrepancies in outbreak timing and magnitude [10, 66]. Additional extensions could involve age-structured transmission, human mobility, and heterogeneous contact patterns [67, 68, 69]. Moreover, our use of a parametric sine function for the infection rate constrains transmission dynamics to a fixed periodic structure. More flexible or nonparametric formulations, such as spline-based or data-driven approaches, could better capture irregular transmission patterns without substantially increasing model complexity. Future studies can also explore hybrid models that combine linear and jump components to capture biologically plausible regimes in which variants with few mutations are followed by those with many more, as observed in chronic SARS-CoV-2 infections [70].

Our modeling framework and insights, though developed in the context of H1N1 outbreaks, are broadly applicable to other influenza viruses with similar antigenic and seasonal characteristics. The interplay between transmission variability, waning immunity, and vaccination coverage is not unique to H1N1 and could help explain resurgence dynamics across different influenza subtypes. In this sense, our approach contributes to a broader understanding of how time-dependent biological and behavioral factors shape the dynamics of respiratory infections. By exploring the effects of environmental variability, immune landscape evolution, and vaccination strategies within a modeling framework, this work enhanced our understanding of large-scale influenza resurgence and may help inform future preparedness strategies.

## Data Availability

All data produced are available online at

http://bit.ly/4n9ymPI

## Acknowledgments

We thank Priscila Born at IOC/FIOCRUZ, Leonardo Bastos, Marcelo Gomes, and the team at PROCC/FIOCRUZ for the insightful comments and support on this project. LMS acknowledges support from the National Science Foundation (NSF DMS-2327844). Some of the computing for this project was performed at the High-Performance Computing Center at Oklahoma State University, supported in part by the National Science Foundation (NSF OAC-1531128).

## Supplementary Material

The supplementary material details the estimation of fixed (time-independent) model parameters, our model fitting algorithm, the parameters obtained from least-squares optimization, a summary of model performances, and a link to the data and code used in this study.

### Time-independent model parameters

#### Population data

To estimate the recruitment rate Λ into the susceptible class, we fitted a linear model to population data from 2009 onward. For each location, the model was initialized with the recorded 2009 population (*N*_0_), and the slope was estimated using the method of least squares, minimizing the sum of squared errors between the model and the observed data up to the year of the large resurgence (see Fig. S1). Overall, population changes were gradual and well approximated by a linear trend across all locations. The estimated growth rates, expressed in week^−1^, are summarized in Table S1.

**Table S1:** Population in 2009 and estimated growth rates.

| Location | Population in 2009 ($N_0$) | Growth rate $\Lambda$ ($\text{week}^{-1}$) |
| --- | --- | --- |
| São Paulo, Brazil | 42, 075, 716 | 137.9351 |
| Colombia | 44, 313, 917 | 186.0822 |
| Croatia | 4, 382, 130 | -5.4751 |
| Iran | 74, 322, 685 | 439.9057 |
| New Zealand | 4, 302, 600 | 14.4896 |
| HHS Region 2 | 28, 0626, 68 | 38.4428 |
| HHS Region 6 | 37, 944, 626 | 194.0172 |
| South Africa | 51, 170, 779 | 356.6660 |
| Turkey | 72, 561, 312 | 343.8197 |

**Figure S1:**
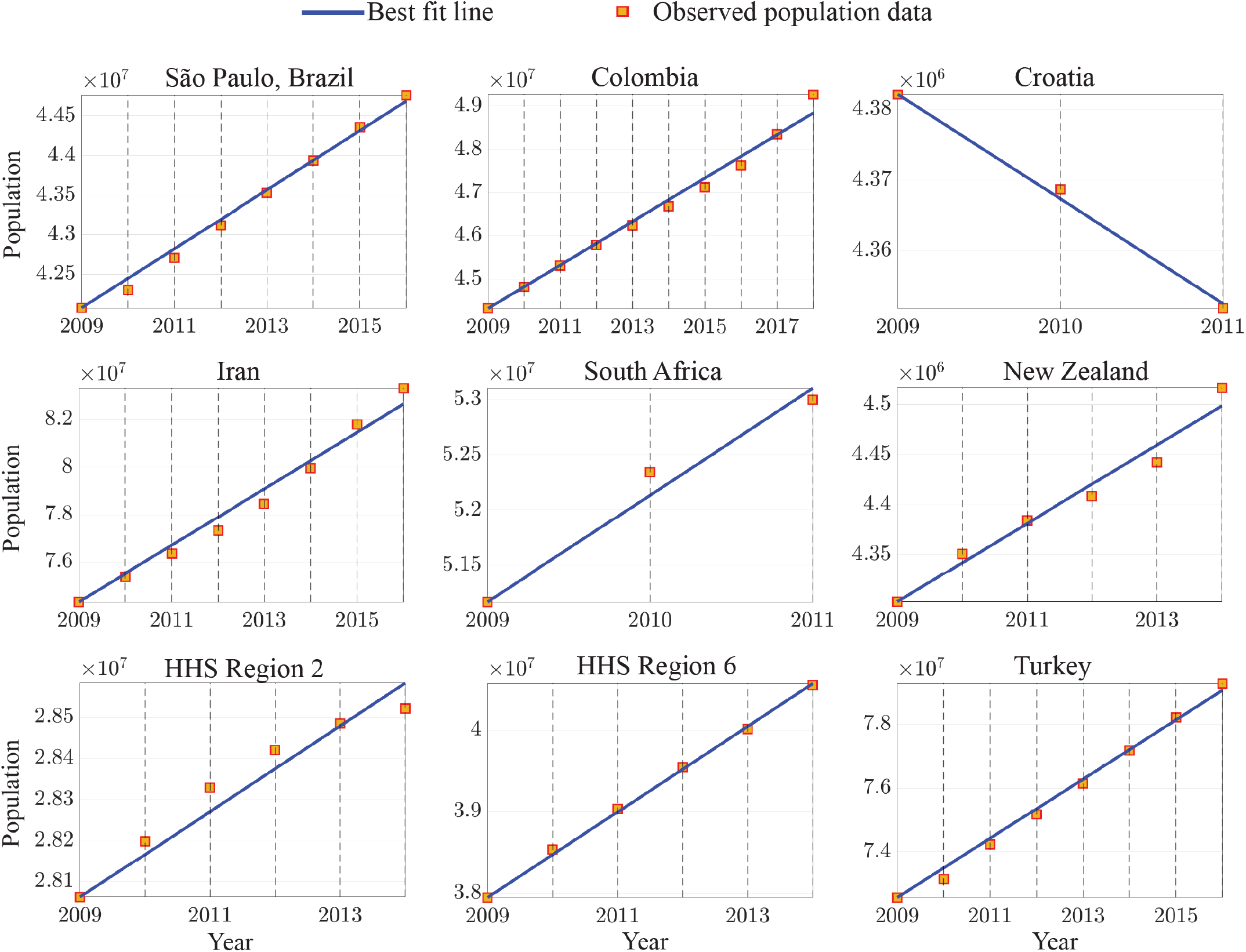
Population data for each location with best-fit linear curve

#### Baseline loss-of-immunity rate (*γ*_0_)

The constant parameter *γ*_0_ represents the baseline rate at which individuals in the recovered class lose immunity. Its reciprocal, 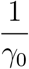, corresponds to the baseline immunity duration, which we use to estimate the loss-of-immunity rate in 2009. Based on influenza studies reporting immunity durations ranging from 4 to 8 years [23], we assumed an average duration of 6 years. Converting to a weekly scale yields

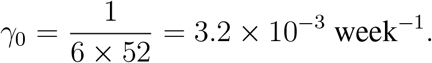

#### Waning rate of vaccine-induced immunity (*ω*) and vaccination periods

The reciprocal 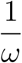 represents the average duration of vaccine-induced immunity for an individual. In this study, we assumed 52 weeks based on [45], leading to

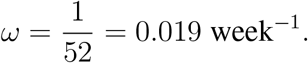

Vaccination periods for each location were estimated based on the months preceding winter and are summarized in Table S2.

**Table S2:**
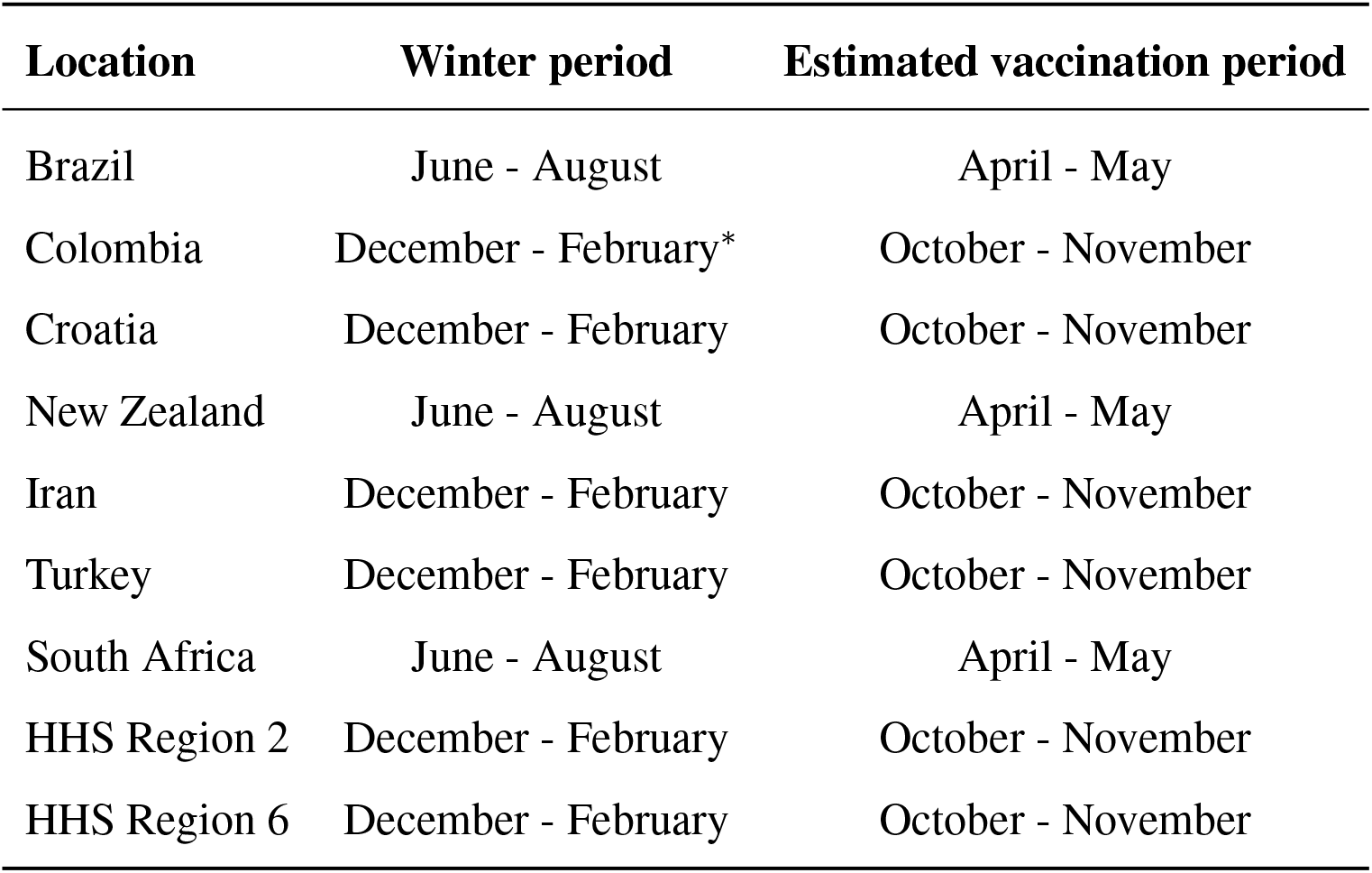
Estimated vaccination periods. Winter periods were defined according to the typical winter months in each region, accounting for their respective hemispheres and climate zones. In Colombia, where a true winter season does not occur due to its tropical climate, the rainy season was considered the equivalent of winter.

| Location | Winter period | Estimated vaccination period |
| --- | --- | --- |
| Brazil | June - August | April - May |
| Colombia | December - February* | October - November |
| Croatia | December - February | October - November |
| New Zealand | June - August | April - May |
| Iran | December - February | October - November |
| Turkey | December - February | October - November |
| South Africa | June - August | April - May |
| HHS Region 2 | December - February | October - November |
| HHS Region 6 | December - February | October - November |

#### Recovery rate (*r*)

The reciprocal 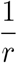 represents the average infectious period, typically ranging from 5 to 7 days [43]. We assumed an average duration of 6 days, which corresponds to 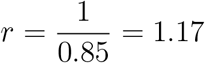 week^−1^.

### Model fitting algorithm

Algorithm (1) describes the process we obtain the best-fit parameter values for a given region with the linear *γ* model. As for the jump model, we modified the algorithm (1) to include the time of the jump of the *γ* function (*τ* ) as the fitted parameter with the lower bound 1 and the upper bound the time span (*T* ) of the fitted region.

#### Algorithm 1

**Grid Search Parameter Optimization for Model Fitting**

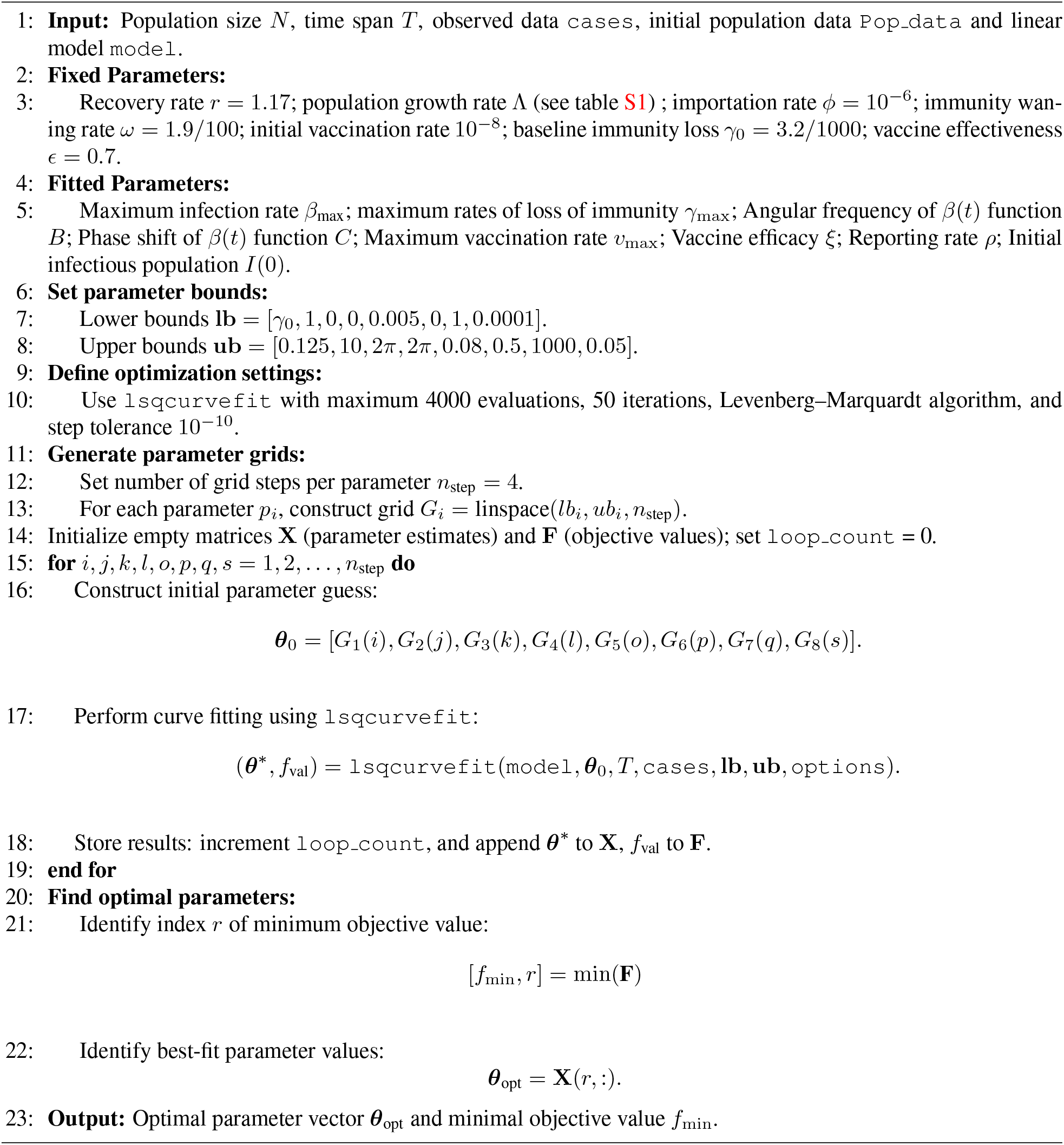

Table S3 displays the percentage of trajectories from the grid search optimization (Algorithm (1)) for which the optimized parameters *θ*^∗^ were close to the global optimum *θ*_opt_. Since the parameter ranges differed across variables, we defined the tolerance for each parameter as one-tenth of its respective range:

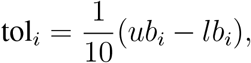

where *ub*_*i*_ and *lb*_*i*_ denote the upper and lower bounds for the *i*-th parameter, respectively. A solution *θ*^∗^ was considered close to *θ*_opt_ if 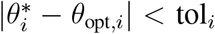 for all parameters *i*. The percentage reported represents the proportion of such solutions relative to the total number of optimization runs. The small percentages suggest that the global optimum occupies a narrow region in parameter space, with most optimization runs converging to local minima.

**Table S3:** Percentage of optimization runs converging to the global optimum.

| Location | Immunity Decay Model | Convergence to optimum (%) |
| --- | --- | --- |
| São Paulo, Brazil | linear | 0.0916% |
|  | jump | 0.0034% |
| Colombia | linear | 0.0122% |
|  | jump | 0.0033% |
| Croatia | linear | 0.0122% |
|  | jump | 0.6944% |
| Iran | linear | 0.0153% |
|  | jump | 0.0034% |
| New Zealand | linear | 0.0168% |
|  | jump | 0.75801% |
| South Africa | linear | 0.009% |
|  | jump | 0.0067% |
| Turkey | linear | 0.15% |
|  | jump | 0.0034% |
| HHS Region 2, USA | linear | 0.0183% |
|  | jump | 0.0060% |
| HHS Region 6, USA | linear | 0.0061% |
|  | jump | 0.0060% |

### Estimated parameters from least-squares optimization and confidence intervals

#### Brazil

**Table S4:** Parameter estimates for São Paulo, Brazil. Best-fit values and 95% CIs obtained from parametric bootstrapping.

| Parameter | Immunity Decay Model | Best-fit value | 95% CI |
| --- | --- | --- | --- |
| $\gamma_{\max}$ | linear | 0.0032 | [0.0032, 0.0032] |
|  | jump | 0.0086 | [0.0075, 0.0094] |
| $\beta_{\max}$ | linear | 2.0598 | [2.0597, 2.0704] |
|  | jump | 2.5160 | [2.5157, 2.5163] |
| Angular frequency of $\beta(t)$ | linear | 0.01976 | [0.0146, 0.0198] |
|  | jump | 0.0295 | [0.0291, 0.03] |
| Phase of $\beta(t)$ | linear | 6.2822 | [6.2818, 6.2832] |
|  | jump | 2.5244 | [2.5242, 2.5248] |
| $v_{\max}$ | linear | 0.005 | [0.005, 0.005] |
|  | jump | 0.005 | [0.005, 0.0099] |
| $\xi$ | linear | 0.5 | [0.5, 0.5] |
| | jump | 0 | [0, $0.05 \times 10^{-4}$] |
| $\rho$ | linear | $2.5466 \times 10^{-4}$ | [$1.360 \times 10^{-4}$, $3.273 \times 10^{-4}$] |
| | jump | $1.3675 \times 10^{-4}$ | [$1 \times 10^{-4}$, $1.6615 \times 10^{-4}$] |
| $\tau$ | jump | 48.8403 | [48.8403, 48.8403] |

#### Colombia

**Table S5:** Parameter estimates for Colombia. Best-fit values and 95% CIs obtained from parametric bootstrapping.

| Parameter | Immunity Decay Model | Best-fit value | 95% CI |
| --- | --- | --- | --- |
| $\gamma_{\max}$ | linear | 0.02 | [0.0170, 0.0242] |
|  | jump | 0.0032 | [0.0032, 0.0032] |
| $\beta_{\max}$ | linear | 10 | [9.998, 10] |
|  | jump | 1.9094 | [1.9094, 1.9095] |
| Angular frequency of $\beta(t)$ | linear | 0 | [0, $1.148 \times 10^{-6}$] |
|  | jump | 6.2768 | [6.2766, 6.2769] |
| Phase shift of $\beta(t)$ | linear | 4.2556 | [4.2556, 4.2556] |
| | jump | 0 | [0, $0.3059 \times 10^{-4}$] |
| $v_{\max}$ | linear | 0.0105 | [0.0100, 0.0116] |
|  | jump | 0.01 | [0.01, 0.01] |
| $\xi$ | linear | 0 | [0, 0.0721] |
|  | jump | 0.5 | [0.5, 0.5] |
| $\rho$ | linear | $1.5227 \times 10^{-4}$ | [$1.294 \times 10^{-4}$, $1.544 \times 10^{-4}$] |
| | jump | $5.5207 \times 10^{-4}$ | [$1.2518 \times 10^{-4}$, $2.0681 \times 10^{-4}$] |
| $\tau$ | jump | 60.3514 | [60.3514, 60.3514] |

#### Croatia

**Table S6:** Parameter estimates for Croatia. Best-fit values and 95% CIs obtained from parametric boot-strapping.

| Parameter | Immunity Decay Model | Best-fit value | 95% CI |
| --- | --- | --- | --- |
| $\gamma_{\max}$ | linear | 0.0257 | [0.0109, 0.0356] |
|  | jump | 0.0100 | [0.0076, 0.0100] |
| $\beta_{\max}$ | linear | 7.05 | [5.7620, 8.4028] |
|  | jump | 4.9944 | [4.7634, 5] |
| Angular frequency of $\beta(t)$ | linear | 0.2398 | [0.2229, 0.2503] |
|  | jump | 0.1290 | [0.1177, 0.1439] |
| Phase shift of $\beta(t)$ | linear | 5.149 | [4.8536, 5.3873] |
|  | jump | 5.8164 | [5.5225, 6.1334] |
| $v_{\max}$ | linear | 0.08 | [0.005, 0.08] |
|  | jump | 0.0134 | [0.005, 0.0248] |
| $\xi$ | linear | 0.5 | [0.2752, 0.5] |
|  | jump | 0.5 | [0.4432, 0.5] |
| $\rho$ | linear | $8.0991 \times 10^{-4}$ | $[5 \times 10^{-4}, 13 \times 10^{-4}]$ |
| | jump | $9.3802 \times 10^{-4}$ | $[6.3501 \times 10^{-4}, 14.835 \times 10^{-4}]$ |
| $\tau$ | jump | 1 | [1, 1.0208] |

#### New Zealanad

**Table S7:** Parameter estimates for New Zealand. Best-fit values and 95% CIs obtained from parametric bootstrapping.

| Parameter | Immuniy Decay Model | Best-fit value | 95% CI |
| --- | --- | --- | --- |
| $\gamma_{\max}$ | linear | 0.0587 | [0.0460, 0.0871] |
|  | jump | 0.01 | [0.0085, 0.01] |
| $\beta_{\max}$ | linear | 3.2591 | [2.7834, 3.7041] |
|  | jump | 3.9148 | 3.4540, 4 |
| Angular frequency of $\beta(t)$ | linear | 0.0459 | [0.0450, 0.0473] |
|  | jump | 0.0233 | 0.0224, 0.0254 |
| Phase shift of $\beta(t)$ | linear | 0.4632 | [0.2947, 0.8597] |
|  | jump | 6.2832 | [6.1373, 6.2832] |
| $v_{\max}$ | linear | 0.005 | [0.005, 0.0390] |
|  | jump | 0.005 | [0.005, 0.0321] |
| $\xi$ | linear | 0.5 | [0, 0.5] |
| | jump | 0.5 | $[8.8846 \times 10^{-8}, 0.5]$ |
| $\rho$ | linear | $2.9337 \times 10^{-4}$ | $[2 \times 10^{-4}, 5 \times 10^{-4}]$ |
| | jump | $3.3975 \times 10^{-4}$ | $[2.1309 \times 10^{-4}, 5.5847 \times 10^{-4}]$ |
| $\tau$ | jump | 1 | [1, 1] |

#### Iran

**Table S8:**
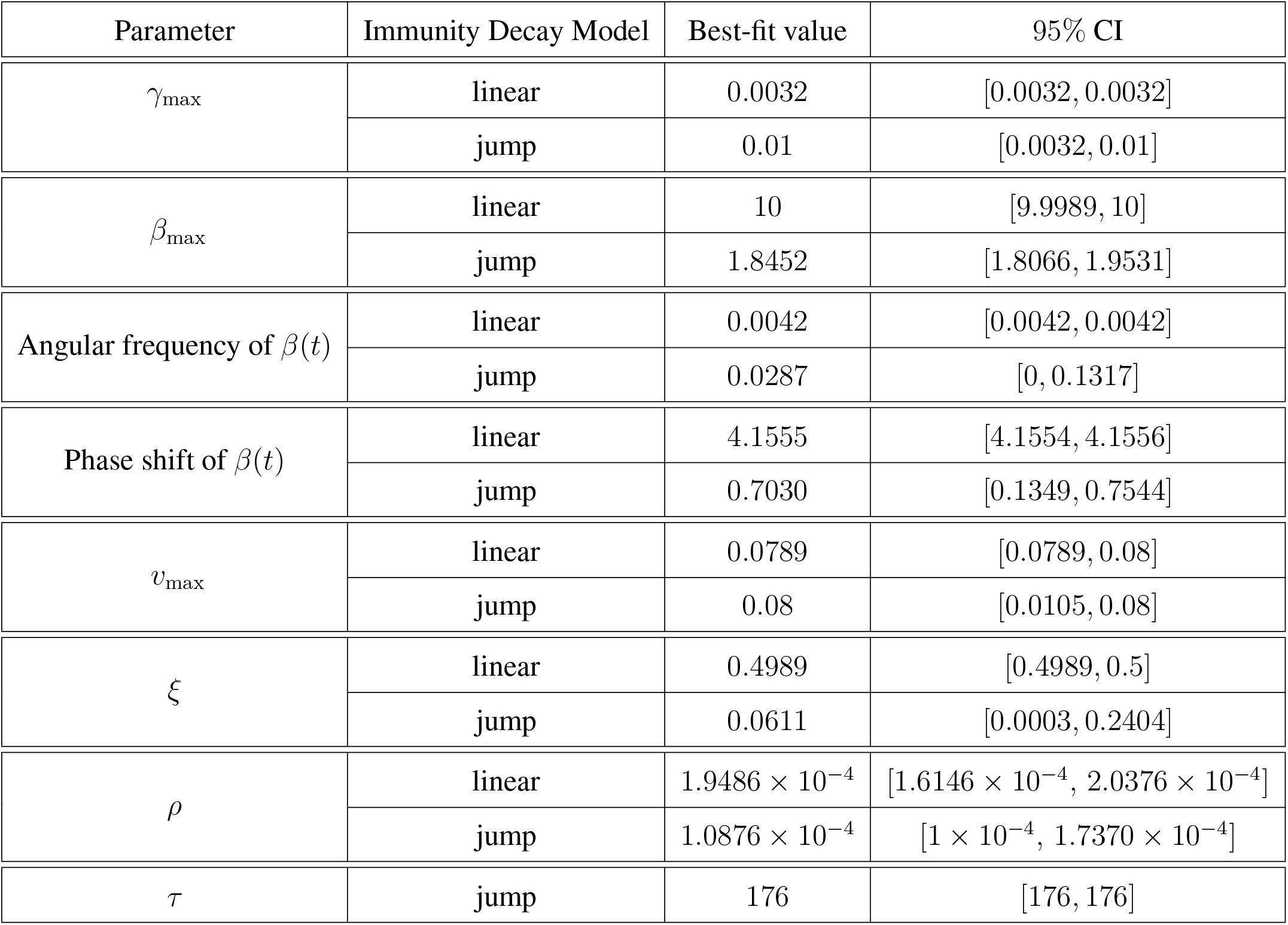
Parameter estimates for Iran. Best-fit values and 95% CIs obtained from parametric bootstrapping.

#### Turkey

**Table S9:** Parameter estimates for Turkey. Best-fit values and 95% CIs obtained from parametric bootstrapping.

| Parameter | Immunity Decay Model | Best-fit value | 95% CI |
| --- | --- | --- | --- |
| $\gamma_{\max}$ | linear | 0.0129 | [0.0032, 0.0195] |
|  | jump | 0.01 | [0.0032, 0.01] |
| $\beta_{\max}$ | linear | 3.8400 | [3.8383, 3.8436] |
|  | jump | 2.8783 | [2.8772, 2.9283] |
| Angular frequency of $\beta(t)$ | linear | 0.0180 | [0.0165, 0.0185] |
|  | jump | 0 | [0, 0.0006] |
| Phase shift of $\beta(t)$ | linear | 6.2828 | [6.2828, 6.2832] |
|  | jump | 0.0534 | [0.0522, 0.0535] |
| $v_{\max}$ | linear | 0.005 | [0.005, 0.0153] |
|  | jump | 0.08 | [0.005, 0.08] |
| $\xi$ | linear | 0.5 | [0, 0.5] |
|  | jump | 0.5 | [0, 0.5] |
| $\rho$ | linear | $1.0556 \times 10^{-4}$ | $[0.6695 \times 10^{-4}, 1.8525 \times 10^{-4}]$ |
| | jump | $1.5917 \times 10^{-4}$ | $[0.8372 \times 10^{-4}, 2.2217 \times 10^{-4}]$ |
| $\tau$ | jump | 221.9301 | [221.9301, 221.9301] |

#### South Africa

**Table S10:**
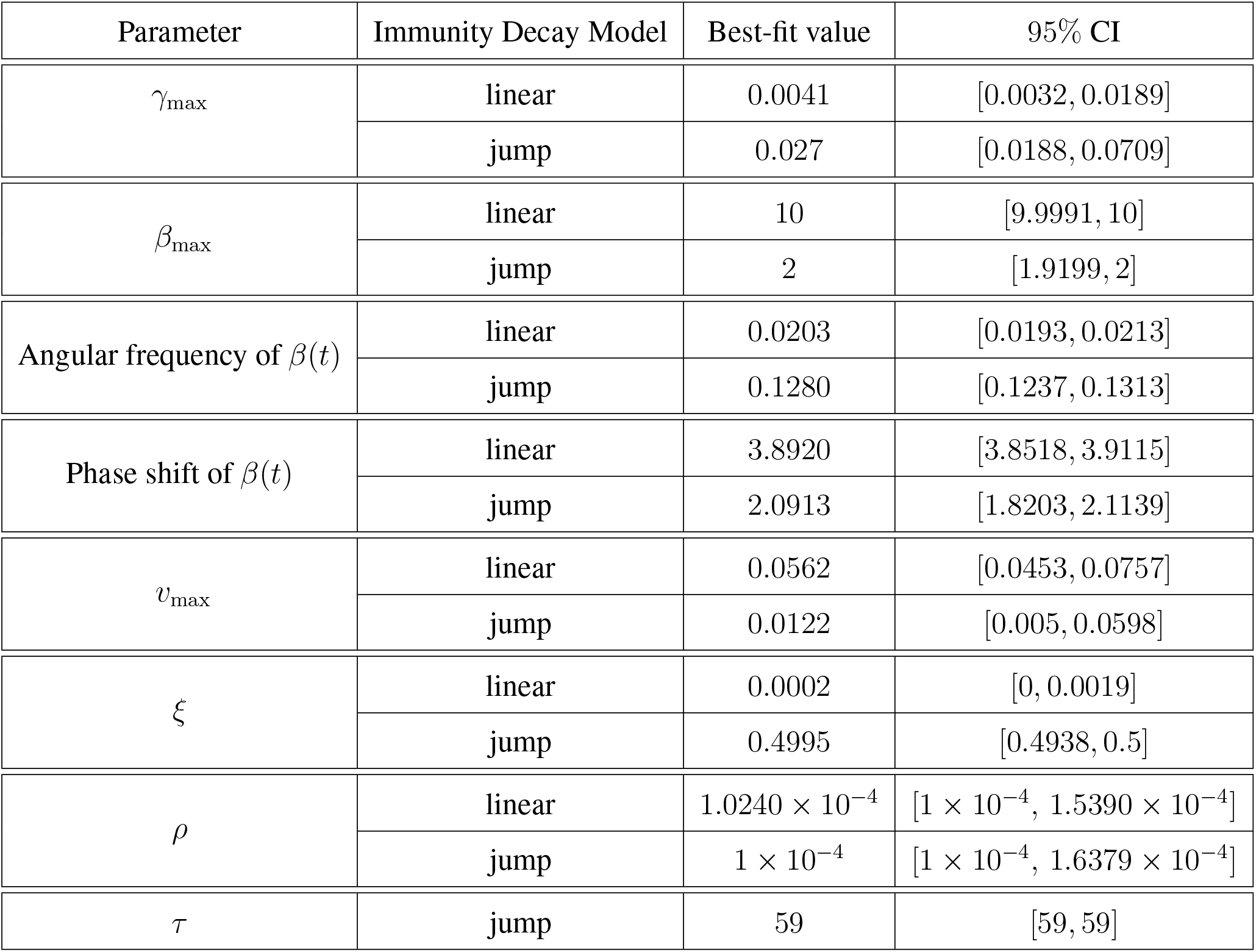
Parameter estimates for South Africa. Best-fit values and 95% CIs obtained from parametric bootstrapping.

#### HHS Region 2

**Table S11:** Parameter estimates for HHS 2 (United States). Best-fit values and 95% CIs obtained from parametric bootstrapping.

| Parameter | Immunity Decay Model | Best-fit value | 95% CI |
| --- | --- | --- | --- |
| $\gamma_{\max}$ | linear | 0.004 | [0.0032, 0.0055] |
|  | jump | 0.0032 | [0.0032, 0.0034] |
| $\beta_{\max}$ | linear | 9.9906 | [9.9769, 10] |
|  | jump | 12.1608 | [11.8924, 12.3074] |
| Angular frequency of $\beta(t)$ | linear | 0.0196 | [0.0120, 0.0199] |
|  | jump | 0.0222 | [0.0215, 0.0240] |
| Phase shift of $\beta(t)$ | linear | 1.8298 | [1.8297, 2.0105] |
|  | jump | 1.2788 | [1.0519, 1.6021] |
| $v_{\max}$ | linear | 0.0541 | [0.005, 0.0701] |
|  | jump | 0.08 | [0.0549, 0.08] |
| $\xi$ | linear | 0.5 | [0.0588, 0.5] |
|  | jump | 0.4977 | [0.1149, 0.5] |
| $\rho$ | linear | $1.8985 \times 10^{-4}$ | $[1.1631 \times 10^{-4}, 2.7578 \times 10^{-4}]$ |
| | jump | $1.7207 \times 10^{-4}$ | $[1.1865 \times 10^{-4}, 2.8047 \times 10^{-4}]$ |
| $\tau$ | jump | 1 | [1, 1.0003] |

#### HHS Region 6

**Table S12:**
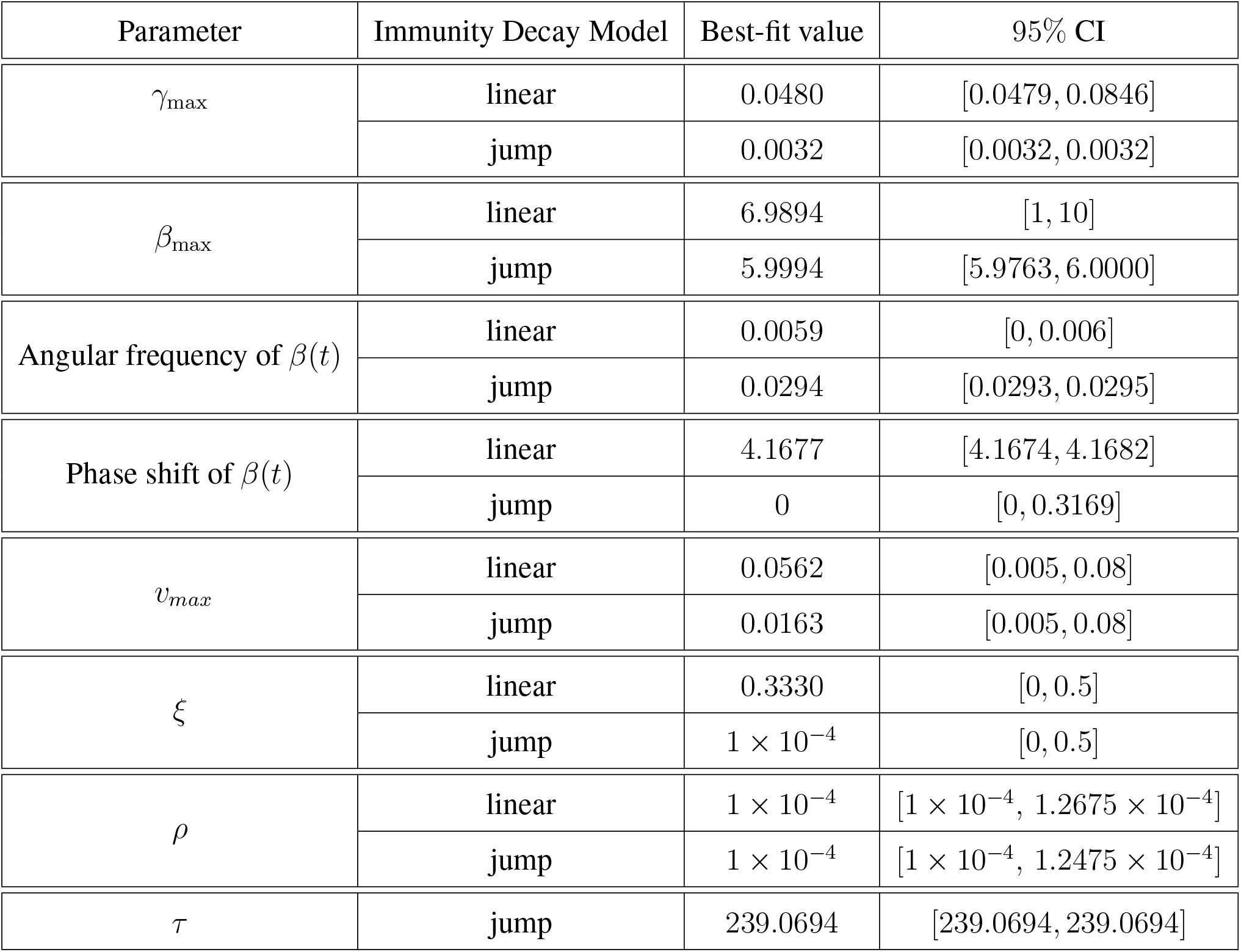
Parameter estimates for HHS 6 (United States). Best-fit values and 95% CIs obtained from parametric bootstrapping.

| Parameter | Immunity Decay Model | Best-fit value | 95% CI |
| --- | --- | --- | --- |
| $\gamma_{\max}$ | linear | 0.0480 | [0.0479, 0.0846] |
|  | jump | 0.0032 | [0.0032, 0.0032] |
| $\beta_{\max}$ | linear | 6.9894 | [1, 10] |
|  | jump | 5.9994 | [5.9763, 6.0000] |
| Angular frequency of $\beta(t)$ | linear | 0.0059 | [0, 0.006] |
|  | jump | 0.0294 | [0.0293, 0.0295] |
| Phase shift of $\beta(t)$ | linear | 4.1677 | [4.1674, 4.1682] |
|  | jump | 0 | [0, 0.3169] |
| $v_{\max}$ | linear | 0.0562 | [0.005, 0.08] |
|  | jump | 0.0163 | [0.005, 0.08] |
| $\xi$ | linear | 0.3330 | [0, 0.5] |
| | jump | $1 \times 10^{-4}$ | [0, 0.5] |
| $\rho$ | linear | $1 \times 10^{-4}$ | $[1 \times 10^{-4}, 1.2675 \times 10^{-4}]$ |
| | jump | $1 \times 10^{-4}$ | $[1 \times 10^{-4}, 1.2475 \times 10^{-4}]$ |
| $\tau$ | jump | 239.0694 | [239.0694, 239.0694] |

### Model Performance

Model performance was evaluated using three different error metrics: Root Mean Square Error (RMSE), Mean Absolute Error (MAE), and symmetric mean absolute percentage error (SMAPE). The formulas for the corresponding metrics are given by

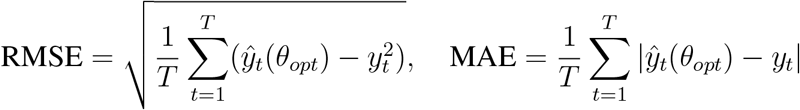

and

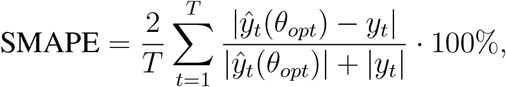

where *ŷ*_*t*_(*θ*_*opt*_) and *y*_*t*_ represent the model output and actual observed cases at time *t*, respectively. Here, *T* denotes the number of weeks. Error values for each location for both models are presented in the Table S13, with the lowest values highlighted in yellow. The linear immunity decay model outperformed the jump immunity decay model on all metrics for São Paulo and HHS Region 2 (USA), and at least two of the three metrics in all other locations, except Turkey, where the jump immunity decay model provided the best performance in terms of RMSE and MAE, although neither model captured the large outbreaks there particularly well. Overall, RMSE and MAE values were relatively low across most locations, indicating good fitting accuracy. Turkey was a notable exception, with much larger errors for both models, suggesting greater difficulty in capturing outbreak dynamics. Even if for most locations the preferred model differed depending on the metric, the differences in error values were small, indicating that both models provided comparable fits.

**Table S13:** Model performance comparison (RMSE, MAE, and SMAPE). Root Mean Square Error (RMSE), Mean Absolute Error (MAE), and Symmetric Mean Absolute Percentage Error (SMAPE) are reported for the linear and jump immunity decay models across all locations. The lowest error for each location is highlighted in yellow.

| Location | Immunity Decay Model | RMSE | MAE | SMAPE |
| --- | --- | --- | --- | --- |
| São Paulo, Brazil | linear | 43.5778 | 13.8190 | 154.0165% |
|  | jump | 47.2714 | 16.7395 | 160.9063% |
| Colombia | linear | 17.5558 | 9.7925 | 139.9145% |
|  | jump | 17.6997 | 9.1485 | 158.9455% |
| Croatia | linear | 22.3348 | 11.1401 | 155.07175% |
|  | jump | 26.3099 | 12.9522 | 124.0109% |
| Iran | linear | 49.7471 | 18.77895 | 180.6155% |
|  | jump | 67.4516 | 15.5259 | 188.0570% |
| New Zealand | linear | 17.6085 | 10.0061 | 157.5454% |
|  | jump | 20.0159 | 10.6950 | 147.7273% |
| South Africa | linear | 14.3537 | 8.3187 | 146.0223% |
|  | jump | 15.9148 | 9.4717 | 142.2827% |
| Turkey | linear | 196.9188 | 63.2264 | 184.2504% |
|  | jump | 193.9137 | 58.9071 | 188.0306% |
| HHS Region 2, USA | linear | 21.3360 | 10.9899 | 166.1053% |
|  | jump | 22.0727 | 11.9082 | 169.7987% |
| HHS Region 6, USA | linear | 77.6263 | 29.6823 | 166.3314% |
|  | jump | 76.2881 | 30.0496 | 170.6639% |

### Data and Codes

All of the data and code are available at the following link.

